# Physician Suicide: A Scoping Review to Highlight Opportunities for Prevention

**DOI:** 10.1101/19004465

**Authors:** Tiffany I. Leung, Sima S. Pendharkar, Chwen-Yuen Angie Chen, Rebecca Snyder

**Affiliations:** Faculty of Health, Medicine, and Life Sciences, Department of Internal Medicine, Maastricht University, Maastricht, The Netherlands; The Brooklyn Hospital Center, Division of Hospital Medicine, Icahn School of Medicine at Mount Sinai, Brooklyn, NY, USA; Department of Primary Care and Population Health, Stanford University, Palo Alto, CA, USA; Library Services, University of Texas Southwestern Medical Center, Dallas, Texas, USA

**Keywords:** Physician Suicide, Burnout, professional, Job satisfaction, Physicians, Suicide, Parasuicide

## Abstract

**Objective:** The aim of this scoping review is to map the current landscape of published research and perspectives on physician suicide. Findings could serve as a roadmap for further investigations and potentially inform efforts to prevent physician suicide.

**Methods:** Ovid MEDLINE, PsycInfo, and Scopus were searched for English-language publications from August 21, 2017 through April 28, 2018. Inclusion criteria were a primary outcome or thesis focused on suicide (including suicide completion, attempts, and thoughts or ideation) among medical students, postgraduate trainees, or attending physicians. Opinion articles were included. Studies that were non-English, or those that only mentioned physician burnout, mental health or substance use disorders were excluded. Data extraction was performed by two authors.

**Results:** The search yielded 1,596 articles, of which 347 articles passed to the full-text review round. The oldest article was an editorial from 1903; 210 (60.3%) articles were published from 2000 to present. Authors originated from 37 countries and 143 (41.2%) were opinion articles. Most discussed were suicide risk factors and culture of practice issues, while least discussed themes included public health and postvention.

**Conclusions:** Consistency and reliability of data and information about physician suicides could be improved. Data limitations partly contribute to these issues. Also, various suicide risk factors for physicians have been explored, and several remain poorly understood. Based on this scoping review, a public health approach, including surveillance and early warning systems, investigations of sentinel cases, and postvention may be impactful next steps in preventing physician deaths by suicide.

## 1 Introduction

Physician suicide is a significant problem for the medical community and general public and is poorly understood, suggesting that important knowledge and implementation gaps towards prevention remain. To address this gap, this literature review aims to describe the state of current knowledge and research on physician suicidal behaviors among medical students, postgraduate trainees, including residents and fellows, and physicians. Suicide is estimated to occur at a higher rate among physicians than the general population and perhaps even other professions (1,2), however, estimates by profession vary (3). Several medical organizations have begun launching various initiatives to address physician well-being, yet efforts to address physician suicide remain organization- or institution-specific. The aim of this scoping literature review is to map the current landscape of published research and perspectives on physician suicide. Findings could serve as a roadmap for informing further study, evidence-based policy, and interventions to prevent physician suicide.

## 2 Methods

This scoping review was conducted following the Arksey and O’Malley framework as expanded upon and outlined within the Joanna Briggs Institute Reviewers’ Manual (4,5). A scoping review differs from other reviews such as a systematic review which gathers and assesses the quality of quantitative evidence to report on the effectiveness of a particular intervention in achieving a certain outcome (6). The research question was broadly designed to gather and analyze articles that mention physician suicide. No date range for the search was specified. The initial searches were performed on August 21, 2017 in Ovid MEDLINE, and October 11, 2017 in Ovid PsycINFO. Authors contributed seed articles that they had previously identified as relevant to physician suicide (2,7–10), which were analyzed by the medical librarian co-author. Search terms and databases were then selected and tested based on this analysis. The search strategy also underwent a peer review process with two additional medical librarians. An updated search was run again on April 28, 2018 in Ovid MEDLINE, PsycINFO, and Scopus. Detailed search terms are available in Appendix 1.

Inclusion criteria were English-language papers with a primary outcome, measure, or thesis focused on death by suicide or suicidal behaviors among physicians, including suicide attempts, and suicidal thoughts and ideation. Physicians included medical students, postgraduate trainees (residents and fellows), and physicians at any career stage. Opinion articles were also included, if they otherwise met inclusion criteria; these included perspectives, letters to the editor or their replies, essays, and viewpoints with a focus specifically on physician suicide. Exclusion criteria were non-English publications and those only pertaining to physician burnout, mental health, substance use disorders, or other media, such as newspaper and magazine articles. Query results totaled 1,596 articles after deduplication. Two authors reviewed abstracts and titles to include in the the full-text review round and disagreements were adjudicated by a third author [initials deleted to maintain integrity of the review process]. Next, during the full-text review round, two authors again reviewed full-text articles for inclusion with a third author adjudicating disagreements. *Covidence*, a literature review management software, was used to review articles during inclusion and exclusion steps

Then, two authors performed data extraction (TL,SP) using a data charting table. Data extracted included: primary thesis or outcome measure (e.g. death by suicide, suicide attempt, suicidal ideation or thoughts), date of publication, authors’ country affiliation, type of publication, study design, country of participants, tools used to ascertain outcome measures, and physician population (specialties, career stage). While reviewing articles, authors used an open coding approach to tag articles by key topics or themes (e.g. suicidal ideation, depression, prevention, substance use, etc.). The aim was to inductively identify key themes across the published literature included in this scoping review. During each subsequent round of review, tags could be added to articles. After all articles were reviewed and tagged by two authors (TL,SP,CYAC), one of the authors (TL) re-reviewed all articles to add tags until a point of saturation was reached and no further topic tags could be added. Tags were then condensed into a core set of themes. These themes were assembled into a framework based on their frequency of occurrence, resulting in a map of the most published themes about physician suicide. Findings are summarized in narrative form.

## 3 Results

The 347 articles that met inclusion criteria (Figure 1) covered a broad range of publication types over time and from countries worldwide. The earliest publications were editorials (Table 1), with the first published in 1903 by an unknown author. Overall, 143 (41.2%) opinion articles were published, suggesting an ongoing public dialogue about physician suicide in academic journals lasting over a century. Of the remaining 204 (58.8%) articles, cross-sectional study design involving a survey was the most commonly used study design. Of these, 13 described interventions intended to prevent physician suicide. Such articles frequently introduced the paper by describing “a tragic case,” when the death of one or more physicians by suicide stimulates the development or implementation of an intervention (11).

**Table 1.** Article characteristics from scoping review of literature on physician suicide, 1903-2018.

| **Publication date range** | **No. (%) of non-opinion articles** | **No. (%) of opinion articles** |
| --- | --- | --- |
| 1900-1919 | 0 | 1 (0.7%) |
| 1920-1949 | 0 | 1 (0.7%) |
| 1950-1959 | 0 | 0 |
| 1960-1969 | 5 (2.4%) | 9 (6.3%) |
| 1970-1979 | 24 (11.7%) | 19 (13.3%) |
| 1980-1989 | 20 (9.8%) | 18 (12.7%) |
| 1990-1999 | 26 (12.7%) | 14 (9.9%) |
| 2000-2009 | 42 (20.5%) | 37 (26.1%) |
| 2010-2018 | 88 (42.9%) | 43 (30.3%) |
| **Study design or publication type** | **No. (%) of non-opinion articles** | **No. (%) of opinion articles** |
| Cross-sectional studies | 85 (41.5%) | N/A |
| Survey-based | 63 |  |
| Review | 34 (16.6%) |  |
| Narrative | 30 |  |
| Case report or case series | 22 (10.7%) |  |
| Case-control | 19 (9.3%) |  |
| Cohort studies | 14 (6.8%) |  |
| Intervention† | 13 (6.3%) |  |
| Other†† | 18 (8.8%) |  |

†Publications about an intervention at minimum describe the intervention.
††Other study designs included: qualitative studies or psychological autopsy (n=8), validation or evaluation of a survey instrument or screening question (3), news coverage (4), poetry (1), a discussion guide (1), or a correction (1).

**Fig. 1.**
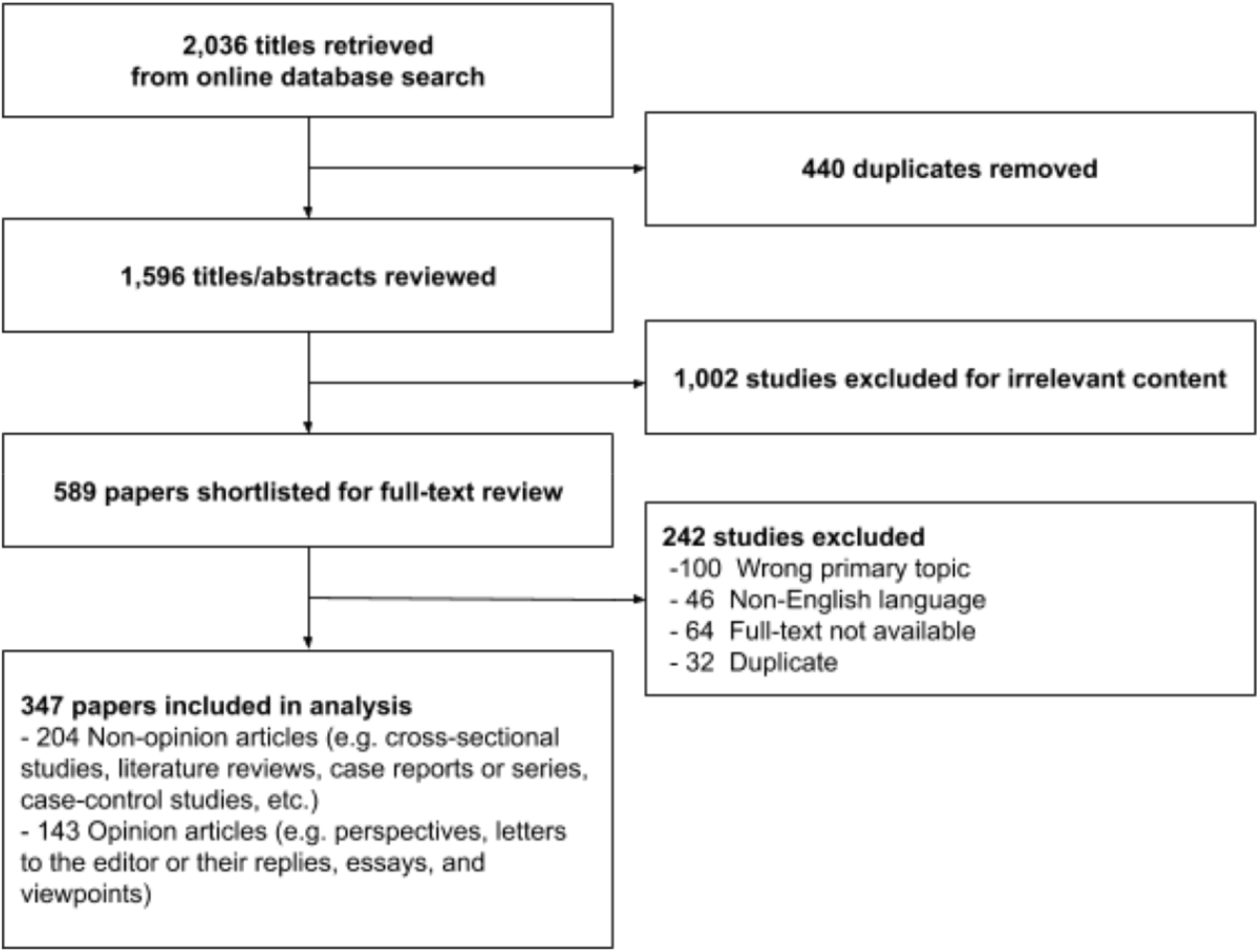
PRISMA diagram of study selection for inclusion.

Authors from 37 countries published articles on physician suicidal behaviors, including: the United States, United Kingdom, Finland, Norway, Sweden, Italy, Japan, Canada, China, Denmark, Hungary, Brazil, Egypt, Germany, Lebanon, Pakistan, Poland, Taiwan, Austria, Belgium, Bosnia and Herzegovina, India, Iran, Malaysia, Nepal, New Zealand, Portugal, Serbia, South Korea, Spain, Thailand, Turkey and the United Arab Emirates. Some countries have only published opinion articles about the subject, including Israel, Nigeria, and Pakistan. Six articles were opinion articles that contained no information about country of origin.

### 3.1 Ascertaining Suicidal Behaviors

#### 3.1.1 Estimating Suicide Incidence

Death by suicide was the only primary topic in 108 (31.1%) of all articles. Non-opinion articles primarily sought to estimate epidemiology of suicidal behaviors among physician populations. In other words, death by suicide was most often studied among attending physicians because of availability of data on occupation and death, particularly in vital statistics or death certificates. However, the tools used to perform such estimates varied. Deaths by suicide were most frequently estimated based on 9^th^ or 10^th^ Revisions of the International Classification of Diseases (ICD-9 or ICD-10) codes on death certificates. Other commonly used sources of data included membership masterfiles from physician organizations or associations (12), published obituaries, and charts from medical records or forensic reports (Table 2). These data are primarily limited by potential undercoding. Undercoding can result from deaths being coded as accidental deaths, for example, leading to underestimation of the actual incidence of suicides among physicians. In an editorial, one author notes, “Suicide is a way to die and not a cause of death. And there are several means to this end: the ICD-10 lists at least 31 different ways to perform a suicide” (13). Rimpela et al articulated this in 1987 also: “Differentiation between suicide, accident, poisoning, and violence as a cause of death is often difficult, and suicide might sometimes be falsely, even deliberately, classified as an accident and even differently in different occupational groups” (14).

**Table 2.** Methods of ascertainment of suicidal behaviors and related risk factors, for 204 non-opinion publications.

|  |
| --- |
| **Methods of ascertainment** |
| **Suicide death** |
| Death certificates (vital statistics by U.S. state or non-U.S. country) |
| Forensic, pathology, or autopsy reports, e.g. from a medical examiner's office |
| Medical chart reviews of physician patients |
| Professional membership masterfiles |
| Obituaries published in journals (e.g. *JAMA*) |
| Family confirmation or interviews |
| **Suicide attempt** |
| Paykel's Instrument (assesses suicidal ideation and suicide attempts) |
| Beck Hopelessness Scale |
| Investigator-developed question(s) or questionnaire |
| **Suicidal thoughts or ideation** |
| Patient Health Questionnaire 9 |
| Beck Scale for Suicidal Ideation |
| Acquired Capability for Suicide Scale |
| Self-harm Behavior Questionnaire |
| Suicidal Behaviors Questionnaire |
| Investigator-developed question(s) or questionnaire |

Suicides are typically reported as a *suicide mortality rate* (SMR) in epidemiologic literature, which is the number of deaths by suicide per 100,000 person-years. A *suicide rate ratio* can be calculated by dividing the SMR among physicians, or a subpopulation thereof, and dividing by a comparison group, such as the general population. Despite these standardized manners of suicide reporting, included papers showed inconsistencies in reporting, often reporting a *crude mortality rate*, which is calculated by the absolute number of physician suicides divided by the number of years in a study period, then reported per 100,000 physicians. For example, in 1968 Craig and Pitts counted 228 suicides among physicians, based on obituary materials collected by the Deaths Editor of the *Journal of the American Medical Association*, between May 1965 and May 1967 (15). Combined with an estimation of approximately 296,000 physicians in the U.S. at the time, this resulted in a *crude annual suicide rate* of 38.4 per 100,000 physicians (15). Multiplying this crude annual suicide rate and the U.S. physician population of approximately 953,000, which was reported by a 2016 census from the Federation of State Medical Boards, leads to an estimate of 366 physician deaths by suicide annually.

Underreporting undermines the accuracy of physician suicide incidence estimates using death certificates, membership files, and medical charts. Surveys also can lead to underreporting. One study surveyed the deans of 116 medical schools in the 1970s about medical student suicides; 88 respondents reported 52 medical student suicides between 1970 and 1978 (16). Another approach to studying physician suicides is psychological autopsy, in which a psychological profile about a person is constructed after their death. Time-intensive and not a standard practice, psychological autopsy involves collecting information through interviews of relatives and healthcare professionals, along with data from forensic examinations, police investigations, psychiatry, medical and social agency records, and any other information available, such as suicide notes (17). Such an approach can elucidate potential contributors to an individual’s death, providing detailed contextual information, their life circumstances, and how they managed such circumstances up until their death. However, psychological autopsies are not systematically applied in the setting of deaths by suicide. Case reports also may describe a psychological profile, in addition to medical and clinical aspects of the case (10,17,18), but this was also infrequently published. In 2013, Gold et al examined data from the U.S. National Violent Death Reporting System (NVDRS), which is a combination of information and data from death certificates, coroner and medical examiner reports, toxicology reports, and law enforcement reports (2), the only study to triangulate multiple data sources to estimate suicide incidence. The wide variety of data sources and methods of estimation suggest that there is no standard.

#### 3.1.2 Suicide Clusters

*Suicide clusters* are suicides that occur near each other, typically with respect to time and geography. *Suicide contagion* refers to spread of information about a suicide via media and other channels, which can increase suicide risk in a community. Only three studies acknowledged these topics. Two opinion papers described suicide clusters that occurred in a short time span and in a focused geographic region, one in Winnipeg, Canada (19) and another cited suicides in Australia that had been reported in news media (20). Both articles focused on intense or distressing working conditions as important factors in the suicide deaths but did not elaborate further on suicide clustering. In fact, the Canadian article, along with one other case series of physicians who died by suicide while on probation in Oregon state (21), used the term “epidemic” rather than “clusters.” Nonetheless, all three papers referred to the concepts of suicide clusters and suggest contagion, even though they do not use these terms explicitly. These were articles were tagged with the public health theme.

Suicide clusters also appeared to follow physician subpopulations, although numbers were small in such case series and opinion articles. Unexpected physician subpopulations included, for example, immigrant physicians (22), physician pilots (23), and physicians who experienced war either as victims or as wartime medics (24,25). In 2016, Bock et al describe a case series of 62 Hungarian Jewish dermatologists during the Holocaust: some emigrated, some died during Nazi rule, and others survived (24). Three of these dermatologists died by suicide, which was “very common among the doctors [under the Nazi ordeal], especially those physicians of the older generation” (24). Another unexpected analysis described the confusing ethical circumstances of physicians as terrorists (26).

#### 3.1.3 Estimating Suicidal Ideation and Attempts

Overall, suicidal ideation was the second most studied thesis among articles included in this literature review, especially suicidal ideation among medical students. To assess suicidal ideation and attempts, surveys were most commonly used; overall, 61 cross-sectional survey studies were performed (Table 1). Validated surveys are available to assess suicidal ideation (Table 2), however, investigator-developed items and surveys were also used. Only four studies compared physician subpopulations between countries. In the Health and Organization among University Hospital Physicians in Europe (HOUPE) study, investigators sought to compare suicidal thoughts and work-related stressors between practicing physicians in Italy and Sweden (27–29). Another study compared lifetime prevalence of suicidal thoughts among practicing physicians in Norway and Germany (30).

Suicide attempts in general were poorly studied in any physician subpopulation, but suicidal behaviors among postgraduate trainees were the least studied of all, with resident physicians’ behaviors studied more than fellows. Among articles reviewed, only two validated tools, Paykel’s Instrument and the Beck Hopelessness Scale, were identified that were specifically designed to assess suicide attempts; otherwise, investigators developed survey items to assess respondents’ self-reported attempts.

### 3.2 Physician-specific Risk Factors

#### 3.2.1 Mental Health and Burnout

Physicians’ risk factors for suicidal behaviors, especially mental health disorders, are among the most common topics of study. Gold et al examined NVDRS data and identified certain risks unique to physicians compared to non-physicians: physicians were more likely to have a job problem preceding death by suicide, and a greater likelihood of known mental health disorders, yet no accompanying increased likelihood of antidepressant therapy (2). Additional risk factors potentially include professional setbacks prior to suicide death, such as facing complaints (21,31), feeling overloaded, working long hours, or being unable to cope with job responsibilities (10), or experiencing disability as a result of medical illness (17,23,32). Related to these conditions, physicians are also at high risk of burnout, which has been found to be associated with suicidal ideation in U.S. medical students (33) and Dutch residents (34), although no direct relationship between burnout and death by suicide has been established. To assess these risk factors, a variety of validated questionnaires were used to inquire about general health, burnout and stress, mental health (e.g. depression, anxiety or insomnia), substance use including alcohol use, and other measures of career, work, personality, and other life experiences (Table 2). Additionally, investigator-developed survey questions were also used, for example, to assess attitudes towards suicidal behaviors of a peer.

Because the earliest publications about physician suicide suggested an increased incidence compared to the general population, certain theories have been applied to attempt to explain the increased suicide risk among physicians. For example, Fink-Miller applied the interpersonal psychological theory of suicidal behavior (IPTS) (35,36) to physician suicide. The IPTS posits three necessary and sufficient precursors to death by suicide: (1) *thwarted belongingness*, a feeling of disconnection with others, (2) *perceived burdensomeness*, a miscalculation that one’s death would relieve burdens on others, and (3) *acquired capability*, habituation to previously provoked fear responses, including losing the fear of pain involved in taking one’s life. This can stem from repeated exposure to painful or provocative stimuli, including events triggering second victimization, such as patients’ poor outcomes, death, and suffering. This can then lead to desensitization when exposed to death in general. *Role strain* describes physician risks as a result of their direct work environment or professional norms; a mismatch between social and institutional norms and the physician’s roles can manifest as an unrealistic expectation of perfect function at a maximum level of competence (37). Professional self-image and identity, along with self-stigma, are also considered relevant mediating themes (17,23,38).

Early studies of suicide rates by specialty suggested that psychiatry and anesthesiology had the highest suicide rates (35). Indeed, these two specialties occurred most frequently in this scoping literature review, followed by surgery or general surgery. However, the findings remain mixed due to data limitations in ascertaining suicide rates as noted previously (39–41). Furthermore, no explanations for the association have been offered other than a proffered but unstudied hypothesis that some medical specialties may possess more acquired capability than others (36,42).

#### 3.2.2 Specialized Knowledge and Access

The *access and knowledge hypothesis* is a commonly discussed risk factor for suicide death among physicians. Physicians acquire specialized medical knowledge of the human body and have access to the means (e.g. prescription drugs) that can cause lethal self-harm. Observational studies of means of suicide death suggest that firearm, prescription drug overdose, and hanging are among the most common methods used by physicians (14,35,41), although this varies by country. Psychological autopsies are consistent with the access and knowledge hypothesis. One set of psychological autopsies in the United Kingdom examined 38 physician suicides, finding that 28 of the physicians died by self-poisoning and 11 by self-injury (other than poisoning), as one physician used both methods (10). In a Finnish psychological autopsy of 7 physician suicides, only 2 had previous suicide attempts, suggesting that physicians may be more likely to cause self-harm leading to death by suicide; in other words, physicians who die by suicide may not have had any prior suicide attempt, which is risk factor in the general population (17). Self-treatment or self-prescription is also a concerning contributor due to specialized access to controlled substances, as “medical doctors exploit their profession for purpose of self-treatment” (17).

#### 3.2.3 Personality Traits, Upbringing, and Cultural Context

Personality and life experience prior to medical training might also contribute to physician suicide risk, although are poorly studied. In a prospective study of Norwegian medical students, the control personality trait, or the degree of compulsiveness, was independently linked to suicidal ideation; and more neuroticism, or the vulnerability personality trait, as a medical student independently predicted more serious suicidal ideations and planning in the first two postgraduate years (9,43,44). Reality weakness, more commonly associated with serious psychiatric pathology, predicted a transition from suicide ideation to planning (9). Neuroticism and tendency towards perfection may even be traits sought in potential medical trainees. One editorial speculates about the influence of adverse childhood experiences on risk for physician suicide (45). One South Korean study found a more than threefold risk of lifetime suicidal ideation, planning, and attempts among medical students who experienced emotional abuse early in life, characterized by “a continuously cold and uncaring parental attitude” (46). In China, medical students were surveyed about parental relationships and parenting communication styles, hypothesizing that this could be a highly influential aspect of the student’s character (47). The study found that for Chinese medical students, a good relationship with parents was statistically significantly associated with less suicidal ideation, plans and attempts.

Some studies sought to identify risk factors unique to their cultural context. Researchers in the United Arab Emirates collaborated with Eskin (48) to again assess medical students’ religiosity and their attitudes towards suicide (49). In this study, investigators suggested that the Islamic faith may serve as a protective factor against suicidal thoughts and death by suicide among their medical students. In Pakistan, a study of suicidal ideation among medical students included religion as a demographic characteristic but did not otherwise include religion in further analyses (50). Among opinion articles, certain conceptualizations of suicide also varied. For example, in one commentary from Japan, two concepts directly link overwork with death: *karoshi* (death due to overwork) and *karojisatsu* (suicide due to overwork) are considered causes of death in the Japanese culture (51) but nowhere else in the world.

#### 3.2.4 Gender

Early studies in the 1970s and earlier explicitly excluded female and minority physicians (43,52,53). Results published in 1999 from the Women Physicians’ Health Study, which surveyed a sample of women physicians from the American Medical Association masterfile (54), described prior studies that reported an odds ratio as high as 4 for women physician suicides compared to other categories of women but that such studies were based on small numbers of suicide deaths. In 2004, Schernhammer performed a systematic review and meta-analysis, concluding that the suicide rate ratio for women physicians was 2.27 compared to the general women population, and 1.41 for male physicians compared to the general male population (1). The 1999 study found that women physicians had similar rates of depression compared to the general population but had lower numbers of suicide attempts (54); the authors postulate that women physicians may have less suicidal intent or a higher rate of completion than the general population, although the study’s scope did not include these outcomes. In 2013, a Polish study of perimenopausal female physicians examined sociodemographic and lifestyle variables’ relationship to participants’ subjective sense of health, including suicidal ideation. Eight of 221 participants reported suicidal thoughts, which was statistically significantly correlated with poorer subjective sense of health (55). Another study in Pakistan appeared to show no gender difference in terms of rates of suicidal ideation among medical students (56). Little data is known on suicide among physicians of gender minorities, as only one perspective piece written by a nurse who wrote about her transgender child who died by suicide (57).

### 3.3 Themes in Physician Suicide Literature

This scoping literature review revealed several themes in physician suicide literature, with greatest attention to identifying risk factors, such as mental health. Culture of practice and context of suicide, which include, for example, mental health stigma, physician attitudes towards suicide, and job conflicts, also were subjects of increased focus. These and the remaining themes identified are illustrated in Figure 2, where the most common themes are positioned at the broadest (top) level of an inverted pyramid. With each successive level of the inverted pyramid, fewer papers describe such issues. We chose an inverted pyramid to illustrate how the weight of the risk factors for physician suicides are balancing precariously over the vertex, where less attention on postvention and public health initiatives for surveillance and prevention. This imbalance quickly identifies potential areas for further work. Themes such as suicide as a public health issue, postvention, and legal issues were far less studied than others.

**Fig. 2.**
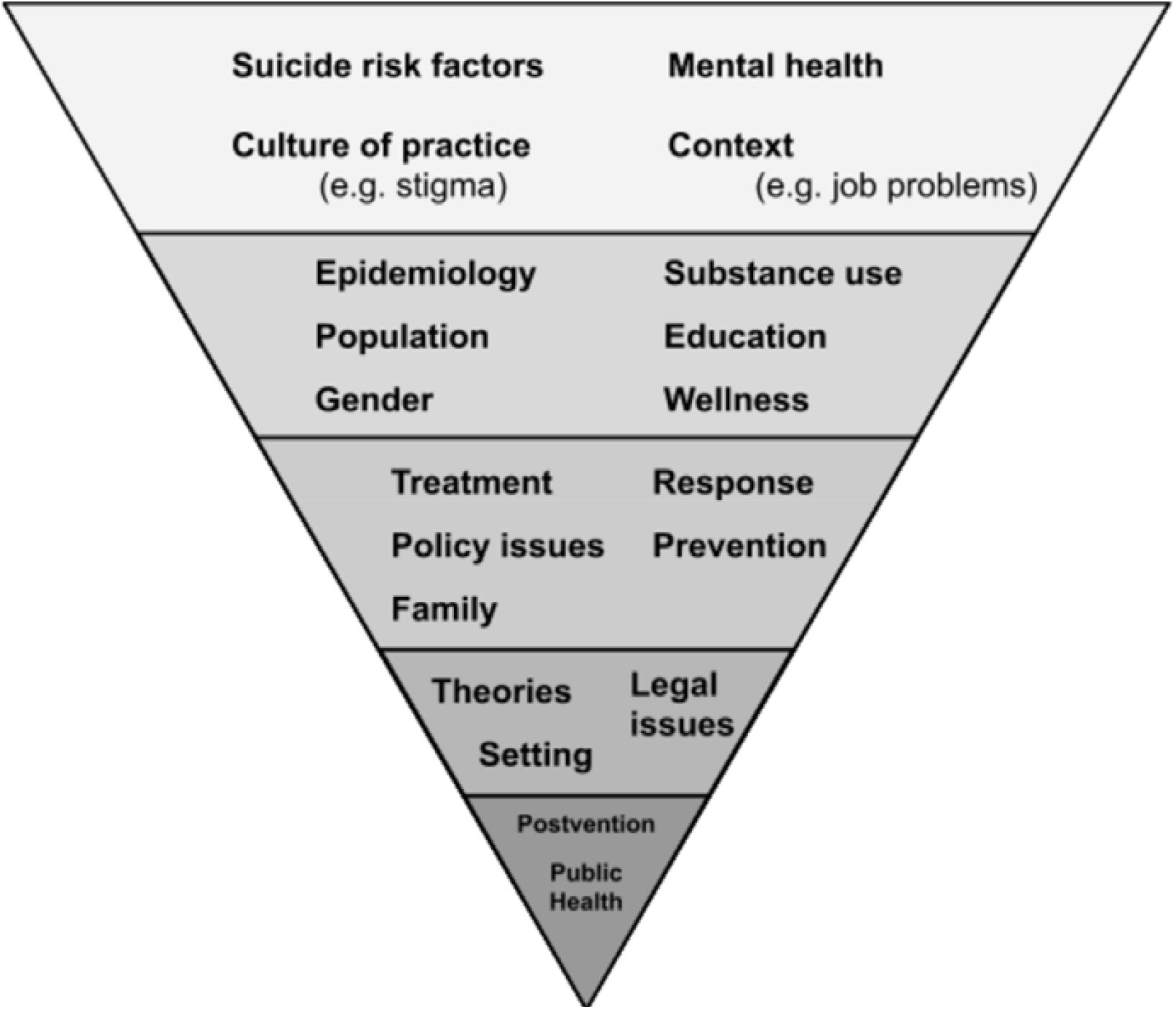
Framework of common publication themes on physician suicide.

In the second level of the framework are epidemiologic and demographic focuses for study, which were previously described in section 3.2. Along with these subjects are a relatively lesser focus on substance use, compared to other risk factors such as mental health, and on developing educational curricula to address knowledge gaps about suicide and its prevention. Wellness is an increasingly popular topic for publication on this level of the framework. In the third level of the framework are prevention approaches. Thirteen articles described interventions with a primary aim of preventing physician suicides. Frequently, programs were implemented as an organizational response to a physician suicide. Five interventions implemented the AFSP’s Interactive Screening Program for suicide prevention (58,59) as a component of a larger initiative to promote physician well-being; three of the five came from the same institution (60–62). Only one of 13 was a randomized controlled trial of a web-based screening program (63).

Treatments and responses to physician suicide were discussed as frequently as prevention; treatments and responses differed because treatments tended to focus on individual physician treatment for suicidal behaviors or related mental health disorders, and responses tended to focus on small group, physician health program, or medical board responses to individual circumstances. For example, Young et al described the difficult grieving process for a healthcare team after the death a physician (64). Patients were told that the physician who had died “was not available,” trust between staff and administrators eroded, morale decreased, and grieving, guilt and other negative consequences remained unaddressed until six weeks after the physicians’ death (64). The process of facilitating a normal grieving process after a suicide death has occurred requires active planning, called *postvention*. In another study by Kaltreider et al, 20 medical students were interviewed after the death of a classmate by suicide (65). This qualitative study found that students experienced intense, intrusive, and persistent thoughts and emotions relating to their classmate’s death, sometimes lasting for months. Proximity to the site of the death, prior exposure to another suicide previously, or familiarity with depression in themselves or family seemed to increase these effects.

Legal and policy issues are increasingly recognized as affecting physician health and well-being, and appear in the third and fourth levels of Figure 2. Physicians may avoid seeking mental health treatment, avoid self-report, and self-treat mental health and other health issues (66,67), out of fear of reputational damage or concern about perceived weakness or impairment (68). Licensure application questions could be improved (68) and compassionate, confidential assistance programs, such as one implemented in the United Kingdom in 2010, could offer bespoke services to physicians who might be considered at risk of suicide (67). In 2015, the UK’s General Medical Council was recommended to fund a pilot program that would include developing a national support service for doctors (69). However, no literature was found to describe such policies in other countries.

Only three articles describe suicide clustering and suicide contagion, which are unique public health concepts pertaining to suicide (19–21). However, no further follow-up or other literature regarding these suicide clusters were found in this scoping review. As a result, the themes of postvention and public health approaches to physician suicide both were at the vertex (bottom) of the inverted pyramid.

## 4 Discussion

As a scoping literature review, this review did not include a quality assessment of quantitative evidence from included papers nor did it perform a meta-analysis or any statistical comparisons of published evidence, which would be characteristic of a systematic review (6). More papers on physician suicide have been published over the last two decades than before. This follows an overall trend in published literature on physician well-being and may also reflect growing public recognition of and decreasing stigma relating to suicide. Furthermore, the proportion of opinion articles published is lower in the recent past compared to previously, indicating growing application of contemporary statistical and scientific methods to the study of physician suicide. Nonetheless, themes identified reveal opportunities for further work. The inverted pyramid framework (Figure 2) suggests that there is a dearth of work on examining physician suicide using typical public health approaches to investigate suicide clusters.

Figure 3 offers a proposed framework that highlights priorities for continued work on physician suicide. This framework highlights the potential for developing an organized public health approach to preventing physician suicide. The pyramid’s base identifies areas for immediate, targeted actions, forming the foundation for the following recommendations for action:

**Fig. 3.**
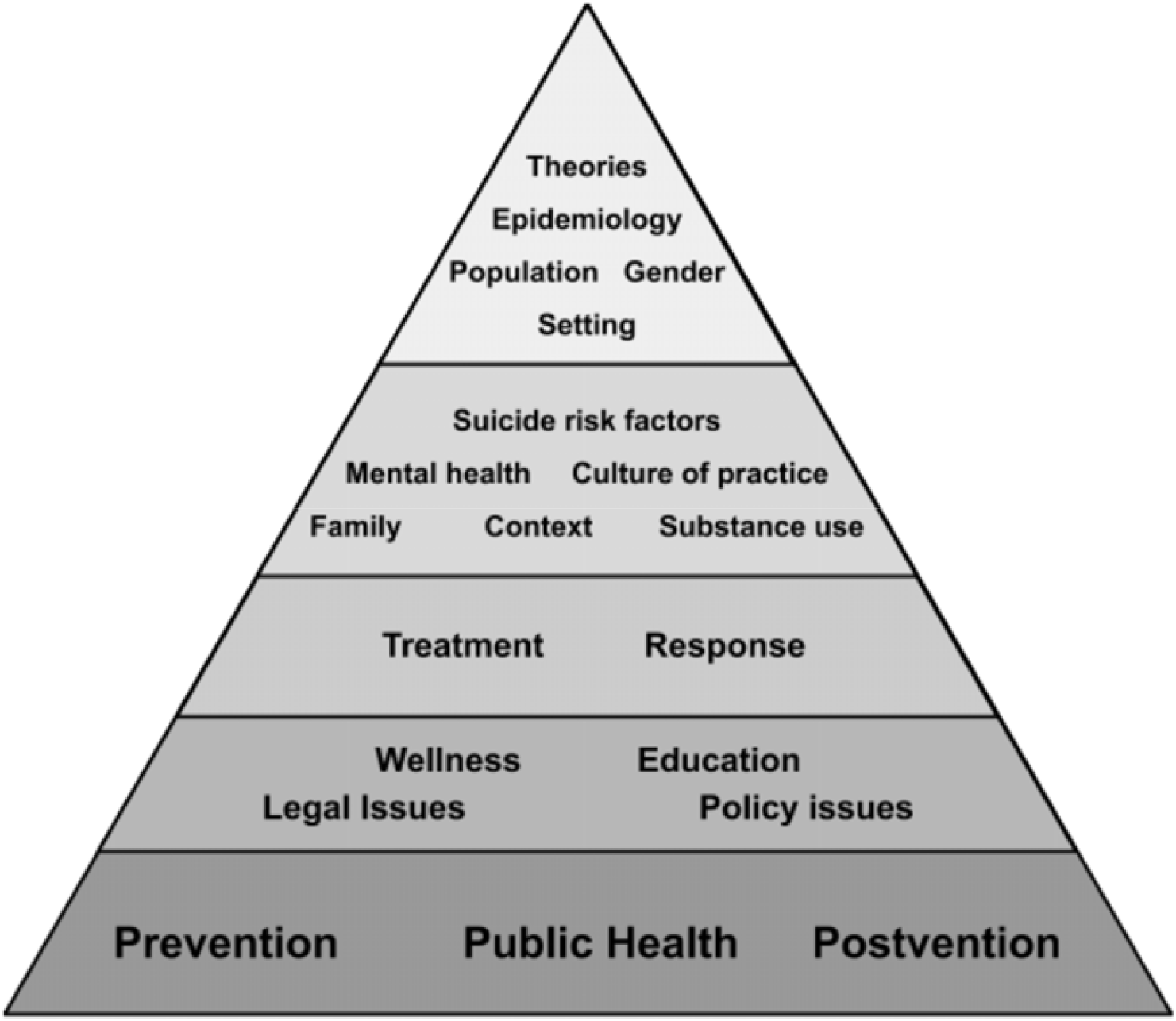
Proposed framework to highlight foundational priorities for continued work on physician suicide.

- Mandatory reporting, an approach used for workplace injury reporting, which could improve surveillance of physician deaths by suicide (prevention);
- Identification of minimum acceptable community standards of validated instruments to assess suicidal ideation and attempts (prevention, public health);
- Development and implementation of evidence-based screening and service provision for suicide risk factors (58–63) (prevention);
- Development of early warning systems, like those used to monitor infectious disease outbreaks, to signal suicide clusters and contagion (public health);
- Implementation and dissemination of best practices, especially regarding postvention or additional directed interventions (postvention);
- Increase education in the healthcare community about suicide warning signs and prevention (education, wellness);
- Standardized education in the medical community about appropriate reporting on suicides to prevent suicide contagion (education);
- Reduction of barriers to help-seeking behavior, reduce systematic stigma in licensing and investigative processes (policy issues, legal issues).

Higher levels in Figure 3 include continued study of persistent risk factors, contextual and cultural challenges, social factors like family, and health inequities resulting from gender, immigrant, or other minority status, among other concerns. Currently available information is insufficient to draw conclusions about physician subgroups or their cultural or health system contexts. Such information on understudied physician subgroups could better inform future revisions of public health, prevention, and postvention programs to include diversity-sensitive suicide prevention approaches.

The strengths of this scoping literature review are its breadth and inclusiveness in examining multiple types of published literature in scholarly journals. However, due to the volume of citations generated by the searches, we excluded grey literature (e.g. conference proceedings, poster presentations, etc.) and snowballing of references was not performed. Additionally, physician suicide involves multiple disciplines, and searching additional electronic databases of peer-reviewed and non-peer-reviewed literature may be appropriate for future updates of such a literature review. Finally, additional articles have been published on physician suicide since the end of the search period of April 2018. Excluding non-English language publications may have led to overrepresentation of countries where English is the official language. Among the 46 non-English language papers excluded (Figure 1), potentially relevant papers were published in German, Spanish, Dutch, Finnish, Japanese, Chinese, French, Swedish, and Hungarian, which could reveal relevant themes in physician suicide, including cultural or health system context. Future studies could identify opportunities for cross-disciplinary and cross-cultural learning and suicide prevention.

## 5 Conclusion

This scoping review offers a landscape view of physician suicide literature and opportunities for further work on physician suicide prevention. Interventions are needed at multiple levels to mitigate the risks of physician suicide, which could begin with an organized public health approach. As a part of such an approach, consistency and reliability of data and information about physician suicides could be improved. Data limitations partly contribute to these issues. For example, annual rates of physician suicide reported and quoted may result from extrapolated estimates from old data, such as those collected by Craig and Pitts in 1968 (15). Reliable and trackable data and information can provide more continuously updated insights into actual suicide mortality rate ratios of physician suicide compared to other populations (70). Also, systems could be developed to better monitor physician suicides, offer early warnings of possible suicide clusters or contagion, and may improve investigation and interventions for the benefit of physicians’ and public health. Physician suicide should be approached as a public health issue and as a shared responsibility between individuals and institutions to prevent physician deaths by suicide.

## Data Availability

Data is available on request from the Corresponding Author.

## 6 Author Contributions

TIL, SP, and CYAC designed the study and performed study selection. RS performed the literature search. TIL and SP extracted data, TIL performed computational analyses, and TIL, SP, and CYAC interpreted the data. TIL drafted the manuscript, except for the Methods section which was drafted by RS, and the entire manuscript was produced with critical revisions and important intellectual content from all authors.

## 7 Acknowledgments

The authors wish to acknowledge Dr. Christine Moutier and Dr. Srijan Sen for their critical review and feedback on the manuscript, as well as Dr. Matthew Goldberg for his early contributions to the background and initial design considerations for this review.

